# Encephalopathies associated with severe COVID-19 present specific neurovascular unit alterations without evidence of strong neuroinflammation

**DOI:** 10.1101/2020.11.01.20217497

**Authors:** Raphaël Bernard-Valnet, Sylvain Perriot, Mathieu Canales, Beatrice Pizzarotti, Leonardo Caranzano, Mayté Castro-Jiménez, Jean-Benoit Epiney, Sergiu Vijiala, Paolo Salvioni Chiabotti, Angelica Anichini, Alexander Salerno, Katia Jaton, Julien Vaucher, Matthieu Perreau, Gilbert Greub, Giuseppe Pantaleo, Renaud Du Pasquier

**Affiliations:** Service of Neurology and Laboratory of Neuroimmunology, Department of Clinical Neurosciences, Lausanne University Hospital (Centre Hospitalier Universitaire Vaudois) and University of Lausanne, Lausanne, Switzerland; Institute of Microbiology, University of Lausanne and University Hospital of Lausanne, Switzerland; Service of Internal Medicine, Department of Medicine, Lausanne University Hospital (Centre Hospitalier Universitaire Vaudois) and University of Lausanne, Lausanne, Switzerland; Service of Immunology and Allergy, Department of Medicine, Lausanne University Hospital and University of Lausanne, Lausanne, Switzerland

## Abstract

**Objective:** Coronavirus disease (COVID-19) has been associated with a large variety of neurological disorders. However the mechanisms underlying these neurological complications remain elusive. In this study we aimed at determining whether neurological symptoms were caused by SARS-CoV-2 direct infection or by either systemic or local pro-inflammatory mediators.

**Methods:** We checked for SARS-CoV-2 RNA by RT-qPCR, SARS-CoV-2-specific antibodies and for 49 cytokines/chemokines/growth factors (by Luminex) in the cerebrospinal fluids (CSF) +/-sera of a cohort of 22 COVID-19 patients with neurological presentation and 55 neurological control patients (inflammatory [IND], non-inflammatory [NIND], multiple sclerosis [MS]).

**Results:** We detected SARS-CoV-2 RNA and virus-specific antibodies in the CSF of 0/22 and 10/21 COVID-19 patients, respectively. Of the four categories of tested patients, the CSF of IND exhibited the highest level of cytokines, chemokines and growth factors. In contrast, COVID-19 patients did not present overall upregulation of inflammatory mediators in the CSF. However, the CSF of patients with severe COVID-19 (ICU patients) exhibited higher concentrations of CCL2, CXCL8, and VEGF-A in the CSF than patients with a milder form of COVID-19. In addition, we could show that intrathecal CXCL8 synthesis was linked to an elevated barrier index and correlated to the increase of peripheral inflammation (serum HGF and CXCL10).

**Conclusion:** Our results point at an absence of massive SARS-CoV-2 infection or inflammation of the central nervous system, but highlight a specific impairment of the neurovascular unit linked to intrathecal production of CXCL8.

## INTRODUCTION

Corona viruses’ outbreaks have been repeatedly associated with neurological disorders. Indeed, human tropic coronaviruses seem able to reach the central nervous system and are found in brain necropsies and in cerebrospinal fluid (CSF) of severe acute respiratory syndrome coronavirus (SARS-CoV) patients ^1, 2^. In mouse model, it has been shown that a strain of human tropic coronavirus is able to reach the olfactory bulb trough the cribriform plate and then to spread through a neuron-to-neuron transmission ^3^.

Coronavirus Disease 2019 (COVID-19) has shown a large range of neurological complications ^4^ that may be classified as followed: critical care-related neurological syndromes either central (encephalopathy with sub-cortical deficit characterized by attention and executive dysfunction) or peripheral (critical care-associated polyneuropathies or myopathies) ^5^; anosmia/dysgueusia ^6^; myelo-meningo-encephalitis ^7^; Guillain-Barré Syndrome (GBS), and its variant affecting cranial nerves (Miller-Fischer Syndrome) ^8^; and cerebrovascular disease (strokes) ^9^.

If data from previous outbreaks point to a neurotropism of the coronaviruses, the pathophysiology underlying SARS-CoV-2-related neurological deficits remains elusive. Indeed, to date, SARS-CoV-2 has only been rarely found in the CSF, suggesting that direct brain infection is unlikely ^10, 11^. Thus, the main hypotheses to explain neurological complications in COVID patients point at mechanisms either related to low grade presence of the virus in the CNS, to disturbance of the blood brain interface by action of peripheral cytokines on brain endothelium or to the presence of an auto-immune response, such as anti-neuronal antibodies by analogy to what occurs in autoimmune encephalitis. However, data firmly establishing one or the other hypothesis are still missing. Yet, the fact that encephalitis/encephalopathies caused by SARS-CoV-2 may respond to corticosteroids ^12, 13^ suggest the involvement of immune mechanisms.

In an attempt to decipher mechanisms underlying neurological symptoms, we looked at SARS-CoV-2-encoding RNA, SARS-CoV-2-specific antibodies and at a panel of 49 cytokines/chemokines/growth factors in the CSF of 77 study patients. Twenty-two of them suffered from COVID-19, and 55 were control SARS-CoV-2-negative patients suffering from inflammatory, including MS or non-inflammatory neurological disorders. We found that COVID-19 patients hospitalized in ICU present signs of blood brain barrier impairment linked to possible astrocyte activation, but no strong immune response in the CSF or obvious CNS infection by the virus.

## MATERIAL AND METHODS

### Study population

All consecutive patients seen at Lausanne University Hospital (CHUV) during the ongoing COVID-19 pandemic (March to end of December 2020) with any neurological manifestations and for whom a lumbar puncture including SARS-CoV-2 PCR on the CSF was performed, were included in this study. Patients with a nasopharyngeal swab positive for SARS-CoV-2 detection by RT-PCR and/or a serology test for anti-SARS-CoV-2 antibodies together with clinical presentation of SARS-CoV-2 infection were defined as COVID-19 patients (25 patients). Of those 25 patients, 3 has a neurological presentation that was likely explained by another condition than COVID-19. Therefore, they were removed from the study. The 22 enrolled COVID-19 patients were further divided into two subgroups whether they requested intensive care unit (ICU), hereafter referred to as severe COVID-19 (13 patients) or not, hereafter referred to as moderate COVID-19 (9 patients). Patients whose nasopharyngeal swab RT-PCR test came back negative were enrolled as controls and classified as inflammatory neurological disorder (IND; 9 patients) or non-inflammatory neurological disorder (NIND; 4 patients) depending on their diagnosis. The control cohort also included 42 patients who were diagnosed with inflammatory neurological disorder (IND; 12 patients), non-inflammatory neurological disorder (NIND; 15 patients) or multiple sclerosis (MS; 15 patients). Serum and CSF of these 42 patients had been drawn prior to the COVID-19 pandemic, in the frame of the COOLIN’BRAIN cohort, between 2009 and 2019. In the subsequent analyses, IND patients who were enrolled during the pandemic period were pooled with the IND patients enrolled before 2020, through COOLIN’BRAIN. The same is true for NIND. Samples (serum and CSF) from MS patients were all obtained during a clinical relapse of MS prior to any corticosteroid treatment.

### Samples

In all study subjects, the lumbar puncture was performed at the acute phase of neurological symptoms. When available, we also analysed serum collected at the same time as the CSF to obtain paired samples (16/22 COVID-19, 14/21 IND, 15/19 NIND, and 15/15 MS patients). Samples of COVID-19 patients and control subjects were run at the same time for detection of SARS-CoV-2 antibodies and cytokines/chemokines/growth factors.

### SARS-CoV-2 PCR

SARS-CoV-2 tests in CSF specimens were performed using our automated platform with an in-house RT-qPCR targeting the E-gene with the primers and probe described by Corman and colleagues ^14^. SARS-CoV-2 tests in respiratory specimens were performed either using our automated platform (at the beginning of the pandemic) or using the cobas SARS-CoV-2 test on the cobas 6800 instrument (Roche, Basel, Switzerland), since March 24, 2020. Both methods were compared and exhibited 99.2% of concordant results ^15^.

### SARS-CoV-2 serology

Anti-SARS-CoV2 IgG specific to the native trimeric Spike (S) protein were quantified using a multiplex bead assay as previously described ^16^. Anti-SARS-CoV2 IgG levels were expressed as a ratio of the mean fluorescent intensity (MFI) signals detected in the serum or CSF samples and the MFI signal detected in the negative control samples. Anti-SARS-CoV2 IgG ratio ≥6 in serum samples were considered positive (threshold set for diagnosis at the Lausanne University Hospital). In the CSF, positivity threshold was set using the mean value of SARS-CoV-2 negative patients + five times SD (positivity threshold = 0.65).

### Cytokine detection by Luminex assay

The concentration of cytokines/chemokines/growth factors in serum and CSF was determined by multiplex bead assay (Thermofisher). The concentration of the following 49 soluble markers was assessed: IL-1alpha, IL-1RA, IL-1beta, IL-2, IL-4, IL-5, IL-6, IL-7, IL-9, IL-10, IL-12p70, IL-13, IL-15, IL-17A, IL-18, IL-21, IL −22, IL-23, IL-27, IL-31, IFN-alpha, IFN-gamma, TNF-alpha, CCL2 (MCP-1), CCL3 (MIP-1α), CCL4 (MIP-1β), CCL5 (RANTES), CCL11 (Eotaxin), CXCL1 (GRO-α / KC), CXCL8 (IL-8), CXCL9 (MIG), CXCL10 (IP-10), CXCL12 (SDF-1α), CXCL13 (BLC), TNF-beta, NGF-beta, BDNF, EGF, HGF FGF-2, LIF, PDGF-BB, PlGF-1, SCF, VEGF-A, VEGF-D, BAFF, GM-CSF and G-CSF. The assay was performed as per manufacturer’s instructions, as previously described ^17^.

### Ethics

This study was approved by the Canton de Vaud Ethical Committee (CER-VD) in the frame of CORO-NEURO study (authorization n° 2020-01123) and COOLIN-BRAIN study (authorization n°2018-01622). All patients included in this study signed specific informed consent.

### Graphical representation and statistical analysis

Graphical representation and statistics were generated using PRISM software (version 8.1.2, GraphPad software, La Jolla, Ca, USA). Multiple group analysis were made using *Kruskal-Wallis test with Dunn’s multiple comparison* test. For analysis with 2 groups, *Mann-Whitney test* was used to determine statistical significance for continuous variables and *Fischer exact test* for categorical variables. Heatmaps and k clustering were made using *ClustVis* web-based tool ^18^.

## RESULTS

### Clinical characteristics

From March to November 2020, 38 patients benefited from a research of SARS-CoV-2 by RT-qPCR and had a concomitant lumbar puncture because of acute neurological symptoms. Of those, 22 patients were diagnosed with COVID-19 based on association of typical symptoms and positive laboratory tests (positive nasopharyngeal swab RT-qPCR (92%) or serology (8%). Their neurological symptoms could not be attributed to another condition than COVID-19 itself. As mentioned in the Methods, there were three additional COVID-19 patients whose neurological disorders were unrelated to COVID-19 (MOG-associated disorders, Lyme’s disease and normal pressure hydrocephalus). In order to avoid any bias, they were not included in further analyses.

Clinical description of SARS-CoV-2 patients is summarized in Table 1. These 22 patients presented with various neurological presentation including encephalopathy (12), encephalitis (2), myelitis (1), optic neuritis (1), Guillain-Barré Syndrome (1), mononeuritis (1) and headache/vertigo (4).

**Table 1.** Detailed clinical and paraclinical characteristics in SARS-CoV-2 infected patient. Abbreviations : EEG (electroencephalogram), MRI (magnetic resonance imaging), FAB (frontal assessment battery), MoCA (Montréal Cognitive Assessment), ICU (intensive care unit), BMI (body mass index), HIV (human immunodeficiency virus). LP (lumbar puncture). *Mann-Whitney test* was used for analysis of continuous variables and *Fischer’s exact test* for categorical ones.

|  | Moderate SARS-CoV-2 (n=9) | Severe SARS-CoV-2 (n=13) | P value |
| --- | --- | --- | --- |
| **Demographic data** |  |  |  |
| • Age, mean [range] | 48,1 [19-69] | 69,1 [53-90] | 0,0059 ** |
| • Sex, Male/Female | 4/5 | 8/5 | 0,67 |
| • BMI, mean [range] | 26,8 [19-37] | 30 [19,5-45] | 0,57 |
| • SARS-CoV-2 mRNA |  |  |  |
| ○ Nasal swab, n (%) | 8 (80) | 12 (100) | - |
| ○ CSF, n (%) | 0 (0) | 0 (0) | - |
| • Pre-existing comorbidities, n (%): |  |  |  |
| ○ Neurodegenerative disease | 0 (0) | 4 (31) | 0,11 |
| ○ Stroke | 2 (20) | 1 (7,7) | 0,54 |
| ○ Immunosuppressive condition/HIV | 0 (0) | 0 (0) | - |
| ○ Malignancy | 0 (0) | 2 (15) | 0,50 |
| **COVID-19 Evolution** |  |  |  |
| • Delay between first symptoms and admission (days), mean [range] | 5,7 [0-21] | 6,9 [0-15] | 0,34 |
| • ICU admission, n (%) | 0 (0) | 12 (100) | - |
| • Mechanical ventilation, n (%) | 0 (0) | 9 (69) | 0,0017 ** |
| • Pulmonary embolism, n (%) | 0 (0) | 1 (7,7) | >0,99 |
| • COVID-19 specific treatments, n (%): |  |  |  |
| ○ Any treatment | 2 (22) | 11 (84) | 0,007 ** |
| ○ Lopinavir | 0 (0) | 4 (30) | 0,11 |
| ○ Tocilizumab | 0 (0) | 1 (7,7) | >0,99 |
| ○ Hydroxychloroquine | 0 (0) | 5 (38,5) | 0,054 |
| ○ Corticosteroids | 2 (20) | 5 (38,5) | 0,65 |
| **Neurological characteristics of SARS-CoV-2 infection** |  |  |  |
| • Main Neurological diagnosis, n (%) |  |  |  |
| ○ Headache/Vertigo | 4 (44) | 0 (0) | 0,017 * |
| ○ Encephalitis/ Meningitis/myelitis | 2 (22) | 1 (7,7) | 0,54 |
| ○ Encephalopathy | 1 (11) | 11 (85) | 0,0015 ** |
| ○ Peripheral nerve | 1 (11) | 1 (7,7) | >0,99 |
| ○ Else | 1 (11) | 0 (0) | 0,41 |
| • Delay in days, mean [range] |  |  |  |
| ○ Between onset of neurological symptoms and LP | 3,3 [0-16] | 6,8 [0-21] | 0,36 |
| ○ Between admission and LP | 4 [0-29] | 22,2 [0-49] | 0,038 * |
| • Modified Rankin Scale, median [range] |  |  |  |
| ○ Before infection | 0 [0-2] | 0 [0-3] | 0,13 |
| ○ At discharge | 1 [0-2] | 3 [2-6] | <0,0001 **** |
| • Brain MRI, n performed: | 10 | 10 |  |
| ○ Diffusion restriction, n (%) | 0 (0) | 1 (7,7) | >0,99 |
| ○ Leptomeningeal/parenchymal Gadolinium enhancement, n (%) | 0 (0) | 1 (7,7) | >0,99 |
| ○ Microbleeds, n (%) | 0 (0) | 1 (7,7) | >0,99 |
| • EEG , n performed: | 2 | 7 |  |
| ○ Encephalopathy, n (%) | 1 (100) | 7 (87,5) | - |
| ○ Irritative, n (%) | 1 (100) | 0 (0) | - |
| • Nerve conduction study, n performed: | 1 | 8 |  |
| ○ Myopathy, n (%) | 0 (0) | 1 (9) | - |
| ○ Polyneuropathy, n (%) | 0 (0) | 6 (54) | - |
| ○ Mononeuritis multiplex, n (%) | 1 (100) | 0 (0) | - |
| ○ Polyradiculopathy, n (%) | 0 (0) | 1 (9) | - |
| • Neuropsychological, n performed: | 0 | 5 |  |
| ○ FAB, median [range] | - | 7 [1-13] | - |
| ○ MoCA, median [range] | - | 15,25 [4-26] | - |

More than half of them (13) were admitted to Intensive Care Unit (ICU), including 9 who required mechanical ventilation. As compared to patients with moderate COVID, patients admitted to ICU were older, had a higher BMI and suffered from more comorbidities. Most of these ICU patients presented with encephalopathy (11/13). Furthermore, some presented signs of critical illness myopathy/polyneuropathy, a diagnosis which was confirmed by nerve conduction studies for most of them (6/8).

No prominent MRI abnormalities were found except in one patient who presented signs of vasculitis including T2 lesions, micro-bleeds and diffusion restriction.

Most electroencephalographic (EEG) recording performed (7/8) showed some abnormalities, consisting mainly in encephalopathic slowing but also irritative activity in one patient with encephalitis.

### CSF characteristics

The CSF of COVID-19 patients was characterised mainly by elevation of protein level and elevated albumin index. Oligoclonal bands were found in a minority of patients. In fact, most of the bands identified on iso-electrofocalisation were shared between the serum and the CSF (type 4; Table 2). These features point toward an opening of the blood brain barrier. Pleocytosis was encountered only in few patients with encephalitic presentation. Interestingly, a striking difference between severe and moderate COVID-19 patients was the higher proportion of elevated barrier index in the former (8/11 −73%) than in the latter (1/6 −16%) (p = 0,049). As expected, the CSF profile was characterized by high protein level and pleocytosis in most IND patients and oligoclonal bands restricted to the CSF (type 2) in all MS patients (Table 2).

**Table 2.** Clinical and CSF characteristics in the tested cohort. Abbreviations: CSF (cerebrospinal fluid). Oligoclonal bands pattern : Type 1: normal CSF; Type 2: oligoclonal IgG restricted to CSF; Type 3: identical IgG in the CSF and the serum with additional IgG bands restricted to the CSF ; Type 4: Identical IgG bands in the CSF and serum; Type 5: Monoclonal IgG bands in CSF and serum (myeloma or monoclonal gammopathy of uncertain significance).

|  | SARS-CoV-2+<br>(n=22) | IND (n=21) | NIND (n=19) | MS (n=15) |
| --- | --- | --- | --- | --- |
| **Demographic data** |  |  |  |  |
| • Age, mean [range] | 60,5 [19-90] | 58 [22-92] | 54 [24-84] | 39 [28-51] |
| **Main Neurological diagnosis, n (%)** |  |  |  |  |
| ○ Headache, n (%) | 4 (18) | 1 (4,8) | 3 (16) | 0 (0) |
| ○ Vascular/Stroke, n (%) | 0 (0) | 1 (4,8) | 2 (10,5) | 0 (0) |
| ○ Encephalitis/ Meningitis, n (%) | 2 (9) | 12 (57) | 0 (0) | 0 (0) |
| ○ Encephalopathy, n (%) | 12 (54) | 2 (9,5) | 0 (0) | 0 (0) |
| ○ Neuro-degenerative disease, n (%) | 0 (0) | 0 (0) | 4 (21) | 0 (0) |
| ○ Myelitis, n (%) | 1 (4,5) | 0 (0) | 0 (0) | 0 (0) |
| ○ Peripheral nerve, n (%) | 2 (9) | 2 (9,5) | 4 (21) | 0 (0) |
| ○ Multiple sclerosis, n (%) | 0 (0) | 0 (0) | 0 (0) | 15 (100) |
| ○ Epilepsy, n (%) | 0 (0) | 0 (0) | 1 (5,3) | 0 (0) |
| ○ Else, n (%) | 1 (4,5) | 1 (4,8) | 5 (26) | 0 (0) |
| **CSF characteristics** |  |  |  |  |
| • CSF proteins |  |  |  |  |
| ○ Level in mg/L, mean [range] | 484 [264-715] | 1479 [309-10588] | 402 [161-895] | 422 [264-594] |
| ○ Elevated (>400 mg/L), n (%) | 13 (59) | 18 (86) | 8 (42) | 8 (53) |
| • Pleocytosis |  |  |  |  |
| ○ Level, mean [range] | 3 [0-21] | 71 [0-540] | 0,95 [0-4] | 15,6 [0-51] |
| ○ Elevated (≥ 5/mm<sup>3</sup>), n (%) | 3 (13) | 13 (61) | 0 (0) | 10 (67) |
| • Elevated Barrier index, n (%) | 9/17 (53) | 15/19 (78,9) | 2/16 (12,5) | 3/15 (20) |
| • Oligoclonal bands pattern, n performed: | 16 | 21 | 14 | 15 |
| ○ Type 1, n (%) | 10 (62) | 8 (38) | 11 (57) | 0 (0) |
| ○ Type 2, n (%) | 0 (0) | 5 (24) | 1 (5) | 9 (60) |
| ○ Type 3, n (%) | 1 (6) | 5 (24) | 0 (0) | 6 (40) |
| ○ Type 4, n (%) | 5 (31) | 0 (0) | 2 (10,5) | 0 (0) |
| ○ Type 5, n (%) | 0 (0) | 1 (4,8) | 0 (0) | 0 (0) |

### SARS-CoV-2 RNA and antibodies detection in the serum and CSF

All 22 COVID-19 patients underwent a RT-qPCR for detection of SARS-CoV-2 RNA in the CSF. It came back negative in all, despite positivity for most (90%) in the nasal swab (table 1). Among patients who had a serological test (14 patients), it came back positive for SARS-CoV-2 antibodies in the blood of 12 (86%). The titers were high (mean ratio of 84.1 for a positivity threshold of ≥6). The absence of anti-SARS-CoV-2 IgG in the remaining two patients, contrasting with a positive nasal RT-qPCR, could be explained by the short time lapse between first symptoms and sampling (respectively 4 and 7 days). Conversely, two patients with IND had SARS-CoV-2-specific antibodies slightly above the positive threshold in the blood (respectively 8.4 and 7.7, positivity threshold ≥6) (Fig. 1C). Of note, these two patients presented with pathologies regularly associated with cross-reactive antibodies: sarcoidosis^19^ and paraneoplastic syndrome ^20^.

**Figure 1.**
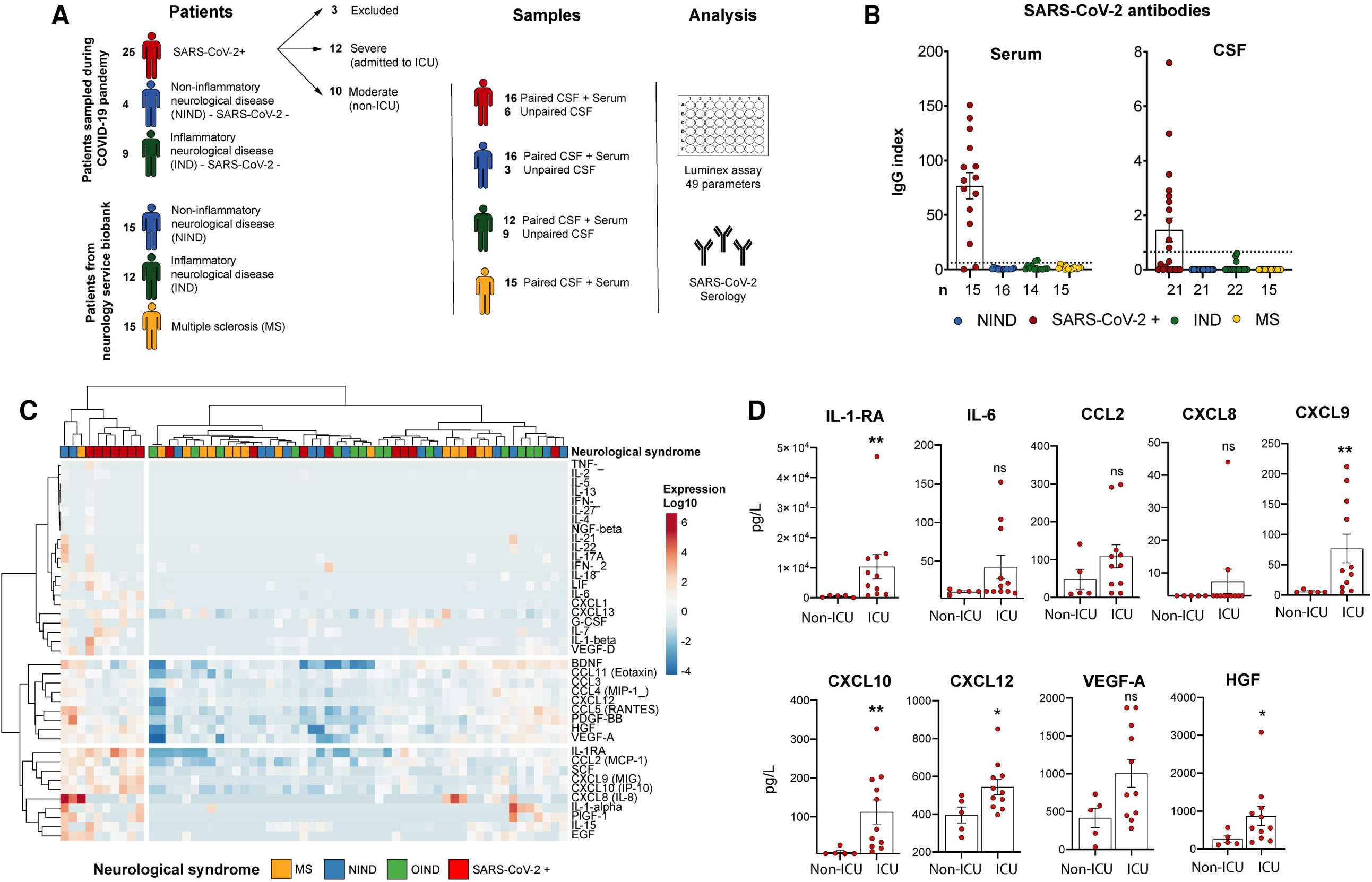
SARS-CoV-2 PCR/serology and Serum Cytokines/Chemokines/Growth factors in COVID-19 patients and control subjects. **(A)** Schematic representation of experimental design. Thirty-eight patients sampled during COVID-19 pandemic and 42 from the biobank of the Service of neurology (COOLIN’BRAIN) were tested for SARS-CoV-2 RNA expression and/or SARS-CoV-2 serology and/or Luminex assay for 49 cytokines/chemokines/growth factors. Of note the CSF +/-serum of all the 42 study subjects who were part of COOLIN’BRAIN was harvested prior to January 2020, thus before the arrival of SARS-CoV2 in Switzerland. **(B)** Antibodies against SARS-CoV-2 detection in the serum and/or the CSF of patient with SARS-CoV-2 infection (red circles, n=14 for serum, n=21 for CSF), IND (green circles, n=14 for serum, n=23 for CSF), NIND (blue circles, n=16 for serum, n=22 for CSF), and MS (yellow circles, n=14 for serum, n=15 for CSF). All 77 study patients were tested for SARS-CoV-2 serologies either in the serum or the CSF or in both compartments. Dotted line represent positivity threshold in the serum. **(C)** Unbiased heat map comparisons of cytokines/chemokines/growth factors within serum of SARS-CoV-2 infected (SARS-CoV-2+) and IND, MS, NIND patients. Expression is represented in log10 scale. K-means clustering was used to determine patients clusters (Cluster 1, n=42; Cluster 2, n=19). Cytokines/chemokines/growth factors with no variations across all patients are not displayed. **(D)** Bar plot representation (mean ± SEM) of IL-1RA, IL-6, CCL2, CXCL8, CXCL9, CXCL10, CXCL12, HGF and VEGF-A expression in the serum of patient with severe (n=11) or moderate (n=5) SARS-CoV-2 infection. Statistical significance was calculated using Mann-Whitney test (ns: not significant, adjusted p:* ≤ 0.05 *** ≤ 0.001, **** ≤ 0.001).

Antibodies against SARS-CoV-2 were detected in the CSF of 10/21 COVID-19 patients with an IgG index >0.65. Conversely, SARS-CoV-2 antibodies were not detected in the CSF of the 55 control study patients (p<0.0001; Fig. 1B).

### Cytokines/Chemokines/Growth factors in the serum and CSF

First, an unbiased analysis of the cytokine panel in the serum of SARS-CoV-2, IND, NIND and MS patients revealed two clusters. Most patients (9/11) with severe COVID-19 belonged to the inflammatory cluster (cluster 2) (Fig. 1C). We found a significant increase of IL-1RA, CXCL9, CXCL10, CXCL12 and HGF in the serum of severe SARS-CoV-2-infected patients as compared to moderate ones (Fig. 1D). In addition, we also found elevated IL-6 levels (>11pg/ml) in 7 out of 11 (63.6%) sera of severe COVID-19 patients versus only 1 out of 5 (20%) of moderate COVID-19 patients (Fig. 1D). These findings recapitulate the inflammatory profile of COVID-19 patients already identified by others ^21, 22^. Similarly, all these cytokines except CXCL12 were also increased in the serum of SARS-CoV-2-infected patients compared to neurological control subjects (MS, IND, NIND) (Fig. e1A).

While SARS-CoV-2-infected patients with neurological conditions were displaying increased inflammatory mediators in the serum, this was not the case in the CSF. Indeed, analyses performed in this compartment revealed that SARS-CoV-2-infected patients were mainly clustered with NIND and MS ones, while the CSF of IND patients exhibited a strong immune signature, characterized by elevated levels of several cytokines, chemokines and growth factors (Fig. 2A). This unbiased clustering was confirmed by individual cytokine/chemokine analysis with significant elevation of IL-1RA, IL-6, CCL4, CCL5, CXCL8, CXCL9, CXCL10, CXCL12, CXCL13 and G-CSF in the CSF of IND as compared to the one of NIND patients (Fig. 2B and data not shown). Even if they did not display as strong an immune signature as IND patients, MS patients showed higher levels of CXCL10, CXCL12, CXCL13 and G-CSF in the CSF (Fig. 2B and data not shown). Of note, COVID-19 patients did not display a significant increase of any of these factors as compared to NIND except for CXCL8 (Data not shown).

**Figure 2.**
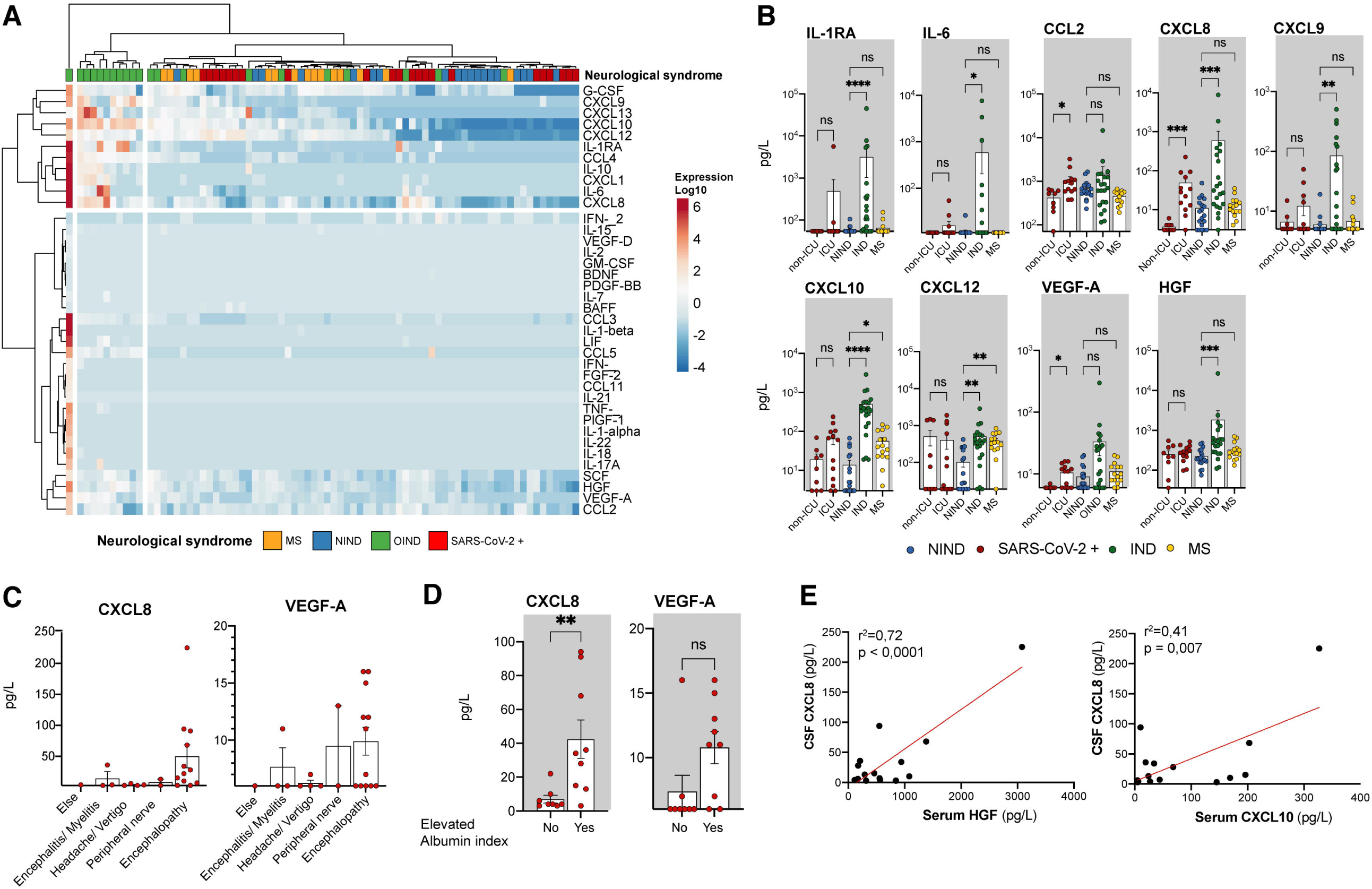
CSF Cytokines/Chemokines/Growth factors in COVID-19 patients. **(A)** Unbiased heat map comparisons of cytokines/chemokines and growth factors within CSF of SARS-CoV-2 infected, and uninfected, i.e IND, NIND and MS patients. Expression is represented in log10 scale. K-means clustering was used to determine patients clusters (Cluster 1, n=1; Cluster 2, n=10; Cluster 3, n=66). Cytokines/chemokines/growth factors with no variations across all patients are not displayed. **(B)** Bar plot representation (mean ± SEM) with log10 scale of IL-1RA, CCL2, CXCL8, CXCL9 CXCL10, CXCL12, HGF and VEGF-A expression in the CSF of patient with SARS-CoV-2 infection (red circles, n=22), IND (green circles, n=21) or NIND (blue circles, n=19) or MS (yellow circles, n=15). Statistical significance calculated using *Kruskal-Wallis test* followed by *Dunn’s multiple comparisons test*, all comparisons performed are displayed (adjusted p:* ≤ 0.05, ** ≤ 0.01, *** ≤ 0.001, **** ≤ 0.001). **(C)** CXCL8 and VEGF-A expression (mean ± SEM) according to neurological diagnosis associated to COVID-19. **(D)** Representation of expression of CXCL8 and VEGF-A in COVD-19 patients with elevated albumin index (n=9) or not (n=8). Statistical significance was calculated using *Mann-Whitney test* (p:* ≤ 0.05, ** ≤ 0.01). **(E)** Linear regression of CXCL8 in the CSF compared to serum HGF (left panel) and serum CXCL10 (right panel).

Accordingly, the clustering based on all inflammatory mediators did not allow to discriminate severe from moderate COVID-19 patients thus demonstrating the absence of cytokine storm in the CSF (Fig. 2A). Yet, interestingly, by individual cytokine/chemokine/growth factors analysis, we found 3 mediators differentially expressed between moderate and severe SARS-CoV-2 patients: CXCL8, CCL2 and VEGF-A (Fig. 2B). Interestingly, the levels of CXCL8 and VEGF-A tended to be higher in the CSF of SARS-CoV-2-infected patients with encephalopathy, 85% of them belonging to the ICU group (Fig. 2C).

As mentioned above, the proportion of patients with an elevated albumin index was significantly higher in severe than in moderate COVID-19 patients. We thus investigated the relationship between this index and the CSF levels of CXCL8, CCL2 and VEGF-A. We observed that CXCL8 was significantly higher in the CSF of patients with elevated albumin index while VEGF-A almost reached statistical significance (p=0.059) but CCL2 showed no difference (Fig. 2D and data not shown). Since an elevated albumin index is a sign of blood brain barrier opening, we next looked at the correlation of CXCL8, VEGF-A and CCL2 levels in serum and CSF. CXCL8 was undetectable in the serum of COVID-19 patients, expect for only 2 of them (Fig. 1D). Serum levels of VEGF-A did not show any correlation with the CSF ones (r^2^=0.004, p value = 0.65), whereas there was a trend for a correlation of CCL2 concentrations between the 2 compartments (r^2^=0.22, p value = 0.06; Fig. e1B). This absence of correlation concentrations between the serum and the CSF, at least for CXCL8 and VEGF-A suggest that the increased CSF levels of these chemokines is not the result of a mere leakage from the periphery, but rather reflects that they are produced intrathecally. Of note, concentrations of CXCL8 and VEGF-A in the CSF correlated together (Fig. e1C), reinforcing the idea that secretion of these factors is linked in COVID-19 context.

Finally, we found that and increased serum level of CXCL10 and HGF was correlated with an increased CSF concentration of CXCL8 (Fig. 2E).

## DISCUSSION

In this study, we attempted to understand the pathogenesis of neurological impairments in the context of COVID-19 disease.

First, we did not detect SARS-CoV-2 RNA in the CSF of any of our 22 patients. Most authors addressing the question also report absence of evidence of viral dissemination in the brain of COVID-19 patients, thus ruling out this causality in the majority of COVID-19-associated neurological disorders. However, we found that about half of SARS-CoV-2-infected patients exhibited virus-specific antibodies in their CSF. Due to the nature of our assay, we were not in a position to determine whether these antibodies in the CSF result from an intra-thecal production or from a passive diffusion through a permeable blood-brain barrier. In any case, it is possible that they are instrumental in the antiviral response in the brain. Supporting these findings, authors recently showed that the CSF from a COVID-19 patient displayed neutralizing antibodies able to prevent neuronal infection in an hiPSC-derived brain organoid model ^23^.

Most importantly, our cohort composed of both severe and moderate patients with paired CSF and serum samples allowed us to study the relationship between peripheral inflammation, neuroinflammation and neurological complications in both sub groups of COVID-19 patients. First, we were able to show that SARS-CoV-2-infected patients, even with a severe disease, do not display a cytokine storm in the CSF despite their sera being the most inflammatory of all 4 groups of study patients. Of note, these patients exhibit an inflammatory profile in the serum similar to what was reported by several groups^21, 22^ thus ruling out that our samples would have been collected after the resolution of the inflammation.

Our findings are in line with a previous report showing low concentrations of IL-6, IL-10 and IFNγ in the CSF of SARS-CoV-2-infected patients with neurological complications ^24^. However, to the best of our knowledge, no other authors have looked so far at such an extensive number of inflammatory mediators, nor have they included such selected control cohorts of study patients with paired CSF/serum samples allowing the assessment of soluble factors in both compartments at the same time.

Interestingly, we could show that patients with a severe COVID-19 exhibited higher CSF levels of CXCL8, CCL2 and VEGF-A than SARS-CoV-2 patients with a moderate disease. In particular, patients suffering from an encephalopathy displayed the highest levels of VEGF-A and CXCL8. Most importantly, despite evidence of blood brain barrier disruption, we found a total absence of correlation between the concentrations of CXCL8 and VEGF-A in the CSF and in the serum. Thus, these data seem to rule out that the increased levels of CXCL8 and VEGF-A in the CSF be a consequence of blood brain barrier opening but rather indicate a local production of these 2 soluble factors in the CNS. Yet, CXCL8 has recently been associated with neurological complications in severe COVID-19^25^.

So, in our study, CXCL8 and VEGF-A were associated with an increased barrier index thus linking the intra-CNS production of these mediators with an involvement of the neurovascular unit (brain endothelium and astrocytes). In the brain, both factors are produced by astrocytes and endothelial cells, and are able to impair the blood brain barrier function^26, 27^. Precisely, microvascular injuries in the brain are a key feature in severe COVID-19 patients^28^.

Following this line of evidence, previous reports have found elevated levels of GFAP and YKL-40 pointing to an enhanced astrocyte reactivity in severe COVID-19 patients with neurological complications. ^25, 29-32^. Yet, when activated, astrocytes can lead to the disruption of the blood brain barrier via both soluble factor production and reduction of astrocytic gap junctions, among other mechanisms. Integrating our findings with what is known in the literature, we propose that in patients with COVID-19 encephalopathy, there is an increased astrocytic reactivity leading to a disruption of the blood brain barrier. Further supporting this hypothesis, this mechanism has been described in patients who present with a delirium associated with critical care ^33^. Furthermore, it is known that hypoxia has strong disrupting effects on the neurovascular unit. Indeed, experimental models have demonstrated that hypoxia causes the production of CXCL8, CCL2 and VEGF-A with discrete production of other inflammatory cytokines^34^ thus mirroring our data from patients. Precisely, hypoxia was present in all patients classified here as severe with 9/13 (69%) requiring mechanical ventilation.

Finally, we found that the intrathecal production of CXCL8 and VEGF-A was correlated to serum HGF and CXCL10. HGF is an instrumental growth factor for tissue repair secreted in response to tissue damage. Its concentration in the serum has been proposed as a prognostic biomarker in severe SARS-CoV-2 infection^22^. In this context, HGF increase seems to appear as a consequence of inflammation and may be a reflection of tissue damage induced by the peripheral cytokine increase.

To conclude, our results suggest that encephalopathies in severe SARS-CoV-2-infected patients are not due to a major viral infection of the brain, neither to a leakage of the cytokine storm from the periphery into the CNS. However, we identify a strong link between encephalopathies and blood brain barrier impairment. Taken together, our study identifies a potential mechanism by which the activation of neurovascular unit cells lead to barrier disruption, as a result of peripheral inflammation and/or hypoxia. The fact that COVID-19-associated encephalopathies respond to corticosteroids support this hypothesis ^12, 13^.

## Data Availability

Data would be stored on CHUV servers but the paper do not refer to supplementary material or external dataset

## ACKNOWLEDGEMENTS

We are grateful to Mrs Géraldine Le Goff for her help in collecting study subjects samples. This work was made possible by grants to RDP from the Swiss National Foundation 320030-179531 and from the Swiss Multiple Sclerosis Foundation.

## AUTHORS CONTRIBUTION

RBV, SP and RDP concepted and designed the study, collected and analyzed the data, drafted the manuscript and figures. MC, BP, LC, MC, JBE, SV, PSC, AA and AS collected the data. KJ, JV, MP GG and GP were involved in conception of the study and analysis of the data.

## Supplemental data

**Fig. e1.**
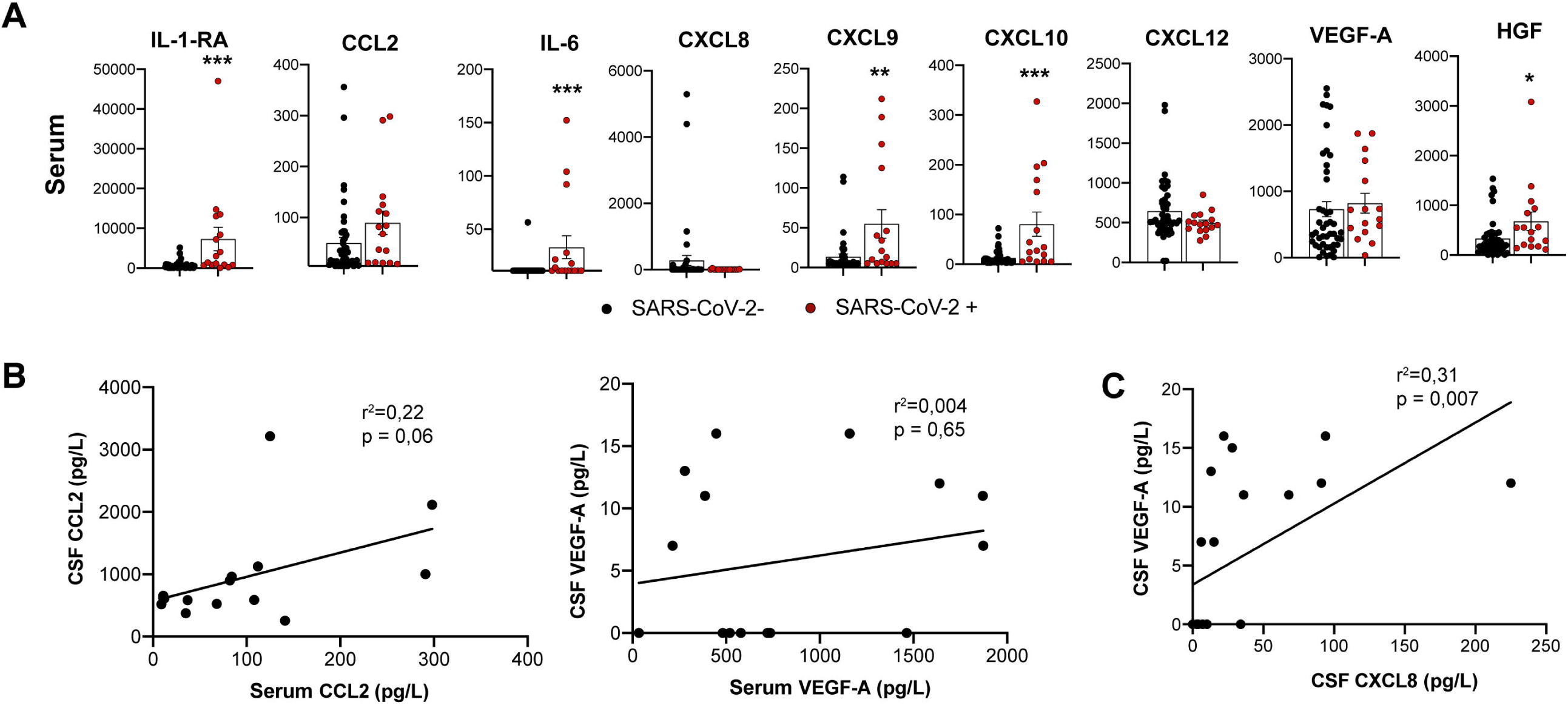
**(A)** Bar plot representation (mean ± SEM) of IL-1RA, IL-6, CCL2, CXCL8, CXCL9, CXCL10, CXCL12, HGF and VEGF-A expression in the serum of patient with (red circles, n=16) or without (black dot, n=45, combination of MS, IND and NIND groups) SARS-CoV-2 infection. Statistical significance was calculated using Mann-Whitney test (p:* ≤ 0.05 *** ≤ 0.001, **** ≤ 0.001). **(B)** Linear regression of level of CCL2 (left panel) and VEGF-A (Right panel) in the CSF and the serum (n=16). **(C)** Linear regression of level of VEGF-A in the CSF in function of CXCL8 expression in the CSF (n=22).

## Appendix 1

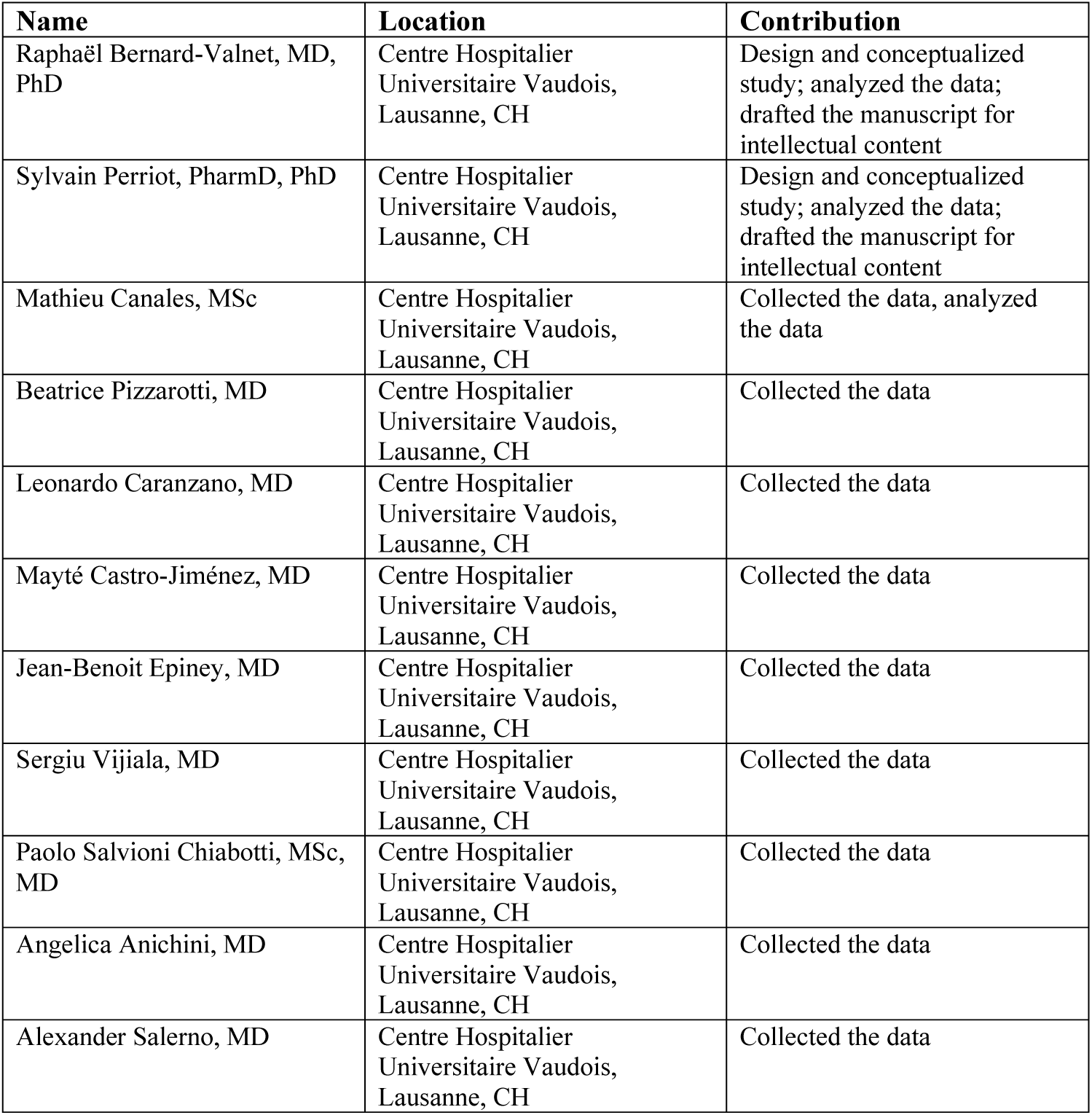
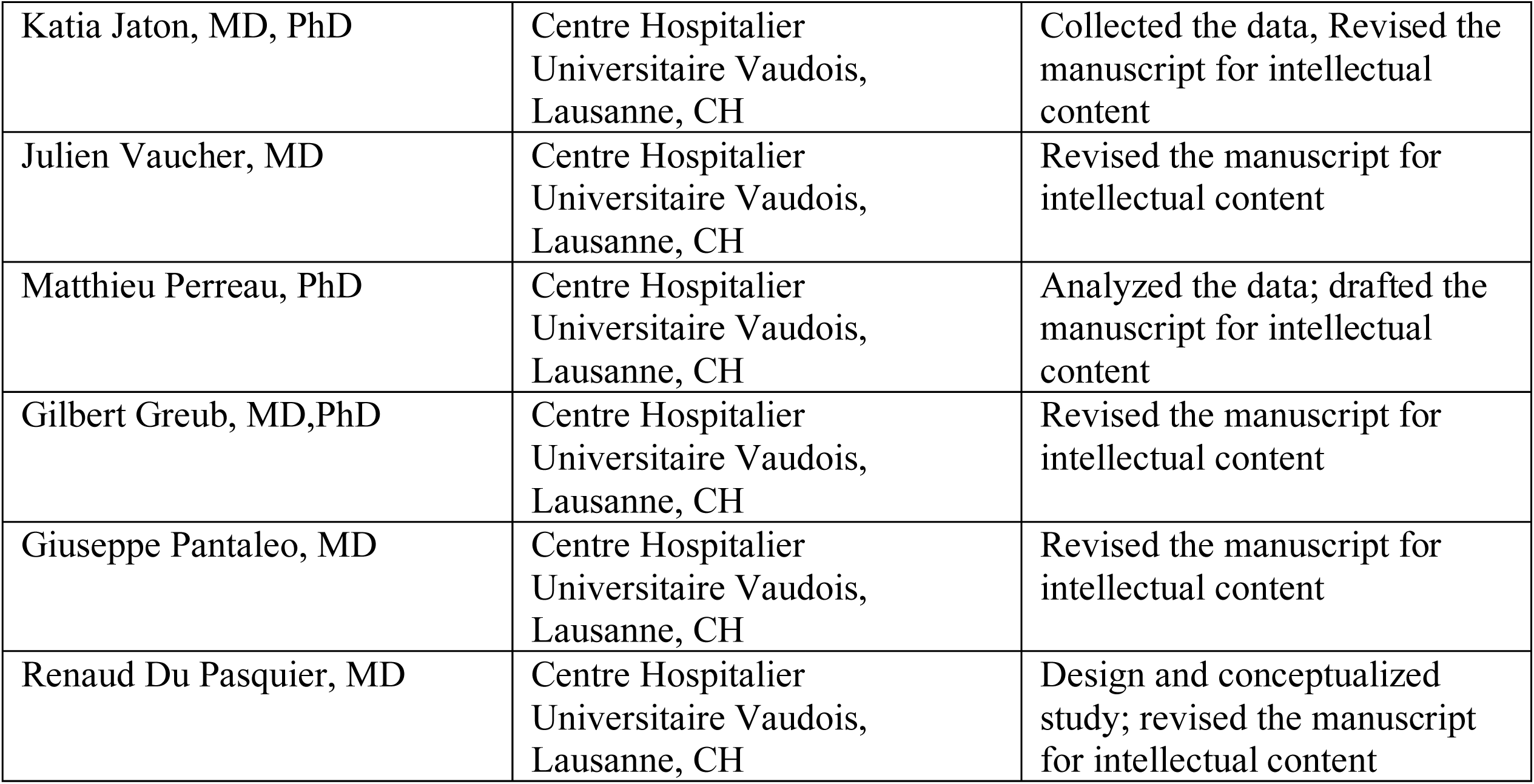

## REFERENCES

1. Gu J, Gong E, Zhang B, et al. Multiple organ infection and the pathogenesis of SARS. J Exp Med 2005;202:415–424.

2. Hung EC, Chim SS, Chan PK, et al. Detection of SARS coronavirus RNA in the cerebrospinal fluid of a patient with severe acute respiratory syndrome. Clin Chem 2003;49:2108–2109.

3. Dube M, Le Coupanec A, Wong AHM, Rini JM, Desforges M, Talbot PJ. Axonal Transport Enables Neuron-to-Neuron Propagation of Human Coronavirus OC43. J Virol 2018;92.

4. Arabi YM, Harthi A, Hussein J, et al. Severe neurologic syndrome associated with Middle East respiratory syndrome corona virus (MERS-CoV). Infection 2015;43:495–501.

5. Helms J, Kremer S, Merdji H, et al. Neurologic Features in Severe SARS-CoV-2 Infection. N Engl J Med 2020.

6. Giacomelli A, Pezzati L, Conti F, et al. Self-reported olfactory and taste disorders in SARS-CoV-2 patients: a cross-sectional study. Clin Infect Dis 2020.

7. Bernard-Valnet R, Pizzarotti B, Anichini A, et al. Two patients with acute meningoencephalitis concomitant with SARS-CoV-2 infection. Eur J Neurol 2020.

8. Toscano G, Palmerini F, Ravaglia S, et al. Guillain-Barre Syndrome Associated with SARS-CoV-2. N Engl J Med 2020.

9. Oxley TJ, Mocco J, Majidi S, et al. Large-Vessel Stroke as a Presenting Feature of Covid-19 in the Young. N Engl J Med 2020.

10. Puelles VG, Lutgehetmann M, Lindenmeyer MT, et al. Multiorgan and Renal Tropism of SARS-CoV-2. N Engl J Med 2020;383:590–592.

11. Virhammar J, Kumlien E, Fallmar D, et al. Acute necrotizing encephalopathy with SARS-CoV-2 RNA confirmed in cerebrospinal fluid. Neurology 2020;95:445–449.

12. Pugin D, Vargas MI, Thieffry C, et al. COVID-19-related encephalopathy responsive to high doses glucocorticoids. Neurology 2020.

13. Cao A, Rohaut B, Le Guennec L, et al. Severe COVID-19-related encephalitis can respond to immunotherapy. Brain 2020;143:e102.

14. Corman VM, Landt O, Kaiser M, et al. Detection of 2019 novel coronavirus (2019- nCoV) by real-time RT-PCR. Euro Surveill 2020;25.

15. Opota O, Brouillet R, Greub G, Jaton K. Comparison of SARS-CoV-2 RT-PCR on a high-throughput molecular diagnostic platform and the cobas SARS-CoV-2 test for the diagnostic of COVID-19 on various clinical samples. Pathog Dis 2020.

16. Fenwick C, Croxatto A, Coste AT, et al. Changes in SARS-CoV-2 Spike versus Nucleoprotein Antibody Responses Impact the Estimates of Infections in Population-Based Seroprevalence Studies. J Virol 2021;95.

17. Perriot S, Mathias A, Perriard G, et al. Human Induced Pluripotent Stem Cell-Derived Astrocytes Are Differentially Activated by Multiple Sclerosis-Associated Cytokines. Stem cell reports 2018;11:1199–1210.

18. Metsalu T, Vilo J. ClustVis: a web tool for visualizing clustering of multivariate data using Principal Component Analysis and heatmap. Nucleic Acids Res 2015;43:W566–570.

19. Grunewald J, Grutters JC, Arkema EV, Saketkoo LA, Moller DR, Muller-Quernheim J. Sarcoidosis. Nat Rev Dis Primers 2019;5:45.

20. Pelosof LC, Gerber DE. Paraneoplastic syndromes: an approach to diagnosis and treatment. Mayo Clin Proc 2010;85:838–854.

21. Del Valle DM, Kim-Schulze S, Huang HH, et al. An inflammatory cytokine signature predicts COVID-19 severity and survival. Nat Med 2020;26:1636–1643.

22. Young BE, Ong SWX, Ng LFP, et al. Viral dynamics and immune correlates of COVID- 19 disease severity. Clin Infect Dis 2020.

23. Song E, Zhang C, Israelow B, et al. Neuroinvasive potential of SARS-CoV-2 revealed in a human brain organoid model. 2020:2020.2006.2025.169946.

24. Helms J, Kremer S, Merdji H, et al. Delirium and encephalopathy in severe COVID-19: a cohort analysis of ICU patients. Crit Care 2020;24:491.

25. Pilotto A, Masciocchi S, Volonghi I, et al. SARS-CoV-2 encephalitis is a cytokine release syndrome: evidences from cerebrospinal fluid analyses. Clin Infect Dis 2021.

26. Haarmann A, Schuhmann MK, Silwedel C, Monoranu CM, Stoll G, Buttmann M. Human Brain Endothelial CXCR2 is Inflammation-Inducible and Mediates CXCL5- and CXCL8-Triggered Paraendothelial Barrier Breakdown. International journal of molecular sciences 2019;20.

27. Shimizu F, Sano Y, Tominaga O, Maeda T, Abe MA, Kanda T. Advanced glycation end-products disrupt the blood-brain barrier by stimulating the release of transforming growth factor-β by pericytes and vascular endothelial growth factor and matrix metalloproteinase-2 by endothelial cells in vitro. Neurobiology of aging 2013;34:1902–1912.

28. Lee MH, Perl DP, Nair G, et al. Microvascular Injury in the Brains of Patients with Covid-19. N Engl J Med 2021;384:481–483.

29. Perrin P, Collongues N, Baloglu S, et al. Cytokine release syndrome-associated encephalopathy in patients with COVID-19. Eur J Neurol 2020.

30. Kanberg N, Ashton NJ, Andersson LM, et al. Neurochemical evidence of astrocytic and neuronal injury commonly found in COVID-19. Neurology 2020.

31. Deigendesch N, Sironi L, Kutza M, et al. Correlates of critical illness-related encephalopathy predominate postmortem COVID-19 neuropathology. Acta Neuropathol 2020.

32. Matschke J, Lutgehetmann M, Hagel C, et al. Neuropathology of patients with COVID- 19 in Germany: a post-mortem case series. Lancet Neurol 2020;19:919–929.

33. Wilson JE, Mart MF, Cunningham C, et al. Delirium. Nat Rev Dis Primers 2020;6:90.

34. Nzou G, Wicks RT, VanOstrand NR, et al. Multicellular 3D Neurovascular Unit Model for Assessing Hypoxia and Neuroinflammation Induced Blood-Brain Barrier Dysfunction. Scientific reports 2020;10:9766.

